# Early-Life Circumstances and Racial Disparities in Cognition among US Older Adults

**DOI:** 10.1101/2021.12.08.21267493

**Authors:** Zhuoer Lin, Justin Ye, Heather Allore, Thomas M. Gill, Xi Chen

## Abstract

**Importance:** Existing studies concentrate on exploring mid-life to late-life risk factors on racial disparities in cognition. Given the critical role of neurocognitive development in early life, understanding contributions of early-life circumstances has important implications for early-stage interventions.

**Objective:** To evaluate the association between early-life circumstances and racial disparities in cognition, and to determine their overall and respective contributions.

**Design, Setting, and Participants:** We assembled three analytic samples from the Health and Retirement Study (HRS) (1995-2018), a nationally representative longitudinal survey of Americans 50 years or older. 17,092 participants, with 13,907 identifying as non-Hispanic White (White) and 3,185 as non-Hispanic Black (Black), were included in the Core sample. The Trauma and PGS samples respectively included 6,533 participants (5,696 White, 837 Black) and 5,532 participants (4,893 White, 639 Black).

**Main Outcomes and Measures:** The main outcomes were cognitive score and cognitive impairment, as assessed by the Telephone Interview for Cognitive Status (TICS). We used the Blinder-Oaxaca Decomposition (BOD) to evaluate disparities in cognitive outcomes between White and Black participants attributable to differences in early-life circumstances.

**Results:** Among all White and Black participants at initial survey, their respective average age were 58.1 (95% CI, 58.0-58.3) years and 55.8 (95% CI, 55.5-56.0) years; their respective average cognitive score were 17.3 (95% CI, 17.2-17.3) points and 14.6 (95% CI, 14.4-14.7) points; and their respective proportion with cognitive impairment were 7.2 (95% CI, 6.8-7.6) percentage points (pp) and 22.9 (95% CI, 21.5-24.4) pp. Across three analytic samples, overall differences in early-life circumstances respectively explained 23.5%–40.4% and 33.8%–65.3% of the racial gaps in cognitive score and proportion of cognitive impairment between White and Black participants. Difference in educational attainment contributed the most. In the Trauma sample, for example, years of education explained 3.1 (95% CI, 1.9-4.3) pp or 18.6% of the racial gap in proportion of cognitive impairment using the baseline assessment, and 3.3 (95% CI, 2.0-4.5) pp or 26.9% using the latest assessment. Additional early-life contributors included educational environments (e.g., ownership of books, parental education, time spent with mothers) and socioeconomic status (e.g., financial difficulty). However, childhood trauma and selected genetic factors were not significant contributors.

**Conclusions and Relevance:** Less favorable early-life circumstances are associated with clinically meaningful and statistically significant racial gaps in cognition.

**Key Points:** *Questions:* How much do differences in early-life circumstances explain late-life disparities in cognitive outcomes between non-Hispanic Black (Black) and non-Hispanic White (White) older adults? What are the key early-life contributors to these racial disparities?

*Findings:* Early-life circumstances contribute substantially to racial disparities in cognitive outcomes over age 50. Educational attainment and early-life educational environment are the most important contributors, even after accounting for a rich set of other early-life socioeconomic, demographic, health, traumatic, and genetic factors.

*Meaning:* Exposure to less favorable early-life circumstances for Black than White adults was associated with large racial gaps in cognitive outcomes.

## Introduction

There are marked differences in rates of cognitive impairment and dementia among the growing US population of older adults. The prevalence rates of cognitive impairment and dementia for non-Hispanic Black (Black) adults are two to three times the rates for non-Hispanic White (White) adults, after accounting for age, sex, education, and late-life comorbidities.^1–3^ The rising proportion of US older adults who are Black or Hispanic may lead to a considerable rise in socioeconomic burden associated with cognitive impairment and dementia.^4–14^ Growing evidence suggests that physical inactivity, smoking, and social isolation are among the strongest risk factors for cognitive impairment and dementia.^15^ Birth cohort trends due to major social, economic, and political changes, as well as medical advances may influence trends of cognitive impairment and dementia.^16^ However, the early-life circumstances through which the racial gap may arise remain under-explored.

Importantly, most research has focused on mid-life to late-life factors; there is little evidence linking factors contributing to neurocognitive development earlier in life with cognitive impairment and dementia in later life. However, brain development is most rapid and plastic early in life.^17, 18^ Strong early-life brain development supports more complex neuritic and intraneuronal connections and cognition, conferring young adulthood and middle age advantage^17^ that may be associated with a more robust cognitive reserve and a lower risk of dementia in later life.^19, 20^

Using a nationally representative longitudinal survey with up to 24 years of follow-up, we investigated how racial gaps in cognitive score and prevalence of cognitive impairment among older Americans are tied to racial differences in early-life circumstances. We tested the following hypotheses: 1) early-life circumstances contribute sizably to racial disparities in cognitive outcomes; and 2) in addition to educational attainment, early-life family educational environments and socioeconomic status are among the most important early-life contributors to cognitive outcomes in old age.

## Methods

### Study Design and Participants

We used the Health and Retirement Study (HRS), a large nationally-representative, longitudinal study of older Americans (aged ≥ 50 years). We assembled a large array of factors on early-life circumstances from four HRS components: the core survey (1995-2018); the Life History Mail Survey (LHMS) (2015, 2017); the Enhanced Face-to-Face (EFTF) Interview (2006-2016); and saliva samples for polygenic scores (PGS) in the EFTF interviews (2006-2012). A description of these data sources is provided in eAppendix A and elsewhere.^21, 22^ The study followed the Strengthening the Reporting of Observational Studies in Epidemiology (STROBE) reporting guideline.^23^

This analysis included HRS participants who had at least one cognitive assessment from 1995 to 2018. Figure 1 provides the sample selection criteria. After excluding participants who did not self-identify as non-Hispanic White (White) or non-Hispanic Black (Black) adults, three analytic samples with increasingly detailed sets of early-life factors were selected, referred to hereafter as the Core, Trauma, and PGS samples, according to the additional specific early-life domains included. The Core sample included 17,092 participants (White: n = 13,907; Black: n = 3,185) and only early-life factors from the core survey; the Trauma sample included 6,533 participants (White: n=5,696; Black: n=837) and additional factors from the EFTF and LHMS; and the PGS sample included 5,532 participants (White: n = 4,893; Black: n = 639) and additional factors from the PGS sample. The Trauma sample was preferred as it included a more comprehensive set of early-life circumstances compared to the Core sample, and larger sample size compared to the PGS sample, which balanced richness of early-life information and sample size among all three samples.

**Figure 1.**
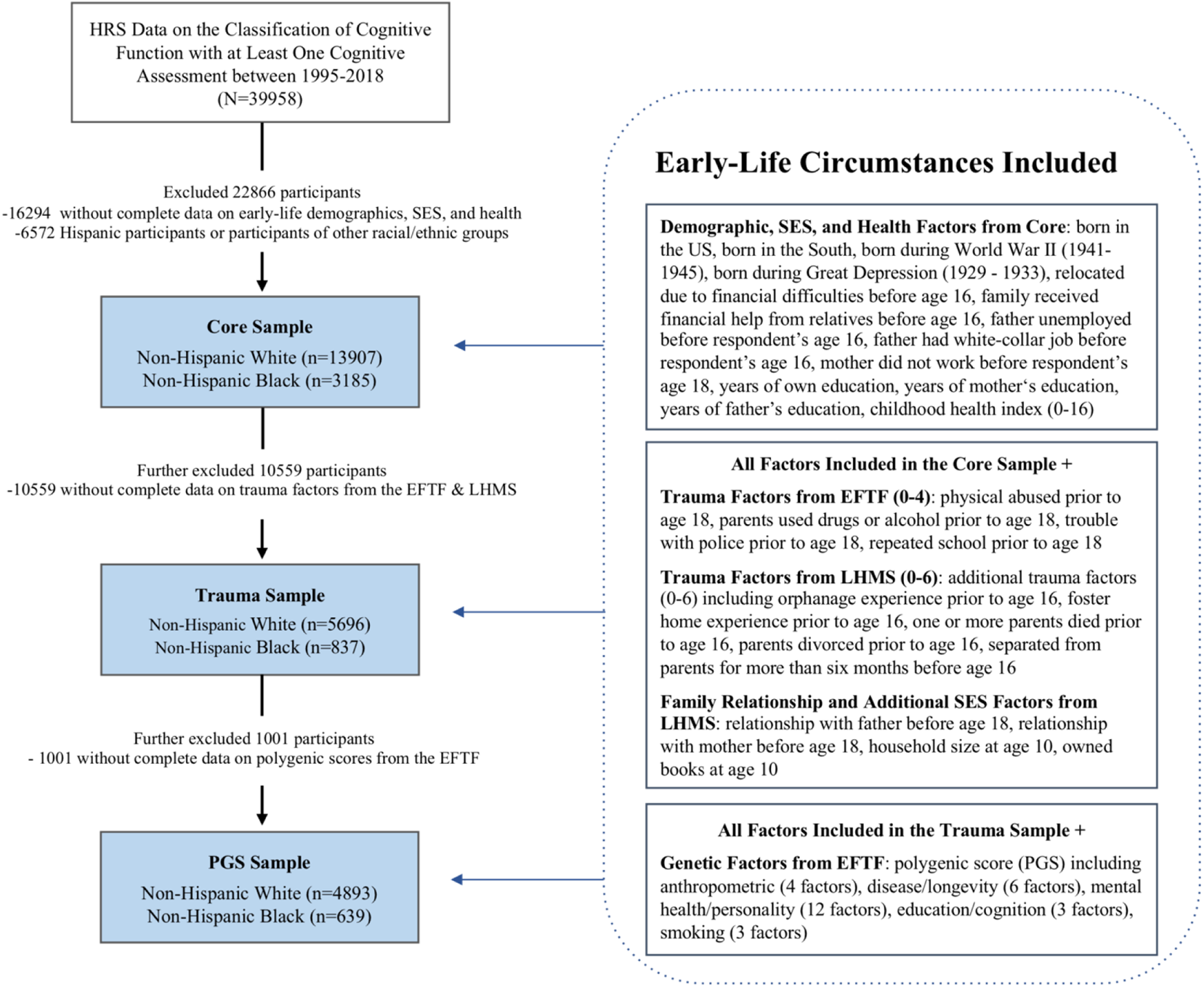
Flow Chart of Sample Selection Process. *Notes*: HRS=Health and Retirement Study; EFTF=Enhanced Face-to-Face (2006-2016); LHMS=Life History Mail Survey (2015, 2017); EFTF and LHMS are separate components conducted within HRS.

For each of the three analytic samples, we analyzed the *earliest* and the *latest* cognitive assessments of the same set of participants, which could be the same assessment for those who were only interviewed once. The time window between the two assessments varied across individuals. This study design is featured by a long duration of follow-up, enabling us to assess the changing role of early-life circumstances over an average of 10 years (see eFigure 1 for the distributions of years gap between the earliest and latest assessment). The HRS study protocol was approved by the Institutional Review Board at the University of Michigan. Data used in this study are de-identified and publicly available.

### Early-life Circumstances

Based on a comprehensive review of the literature, we selected and classified a wide spectrum of early-life factors that may contribute to cognitive impairment, encompassing early-life demographics, socioeconomic status (SES), health, trauma, familial relationships, and genetics. As shown in Figure 1, the Core sample included 13 demographics, SES, and health events (a 16-point index) factors from the HRS core survey. The Trauma sample further included a 4-point trauma index from the EFTF, an additional 6-point trauma index from the LHMS, as well as family relationship and additional SES factors from the LHMS. Lastly, polygenic scores related to anthropometrics, diseases and longevity, mental health and personality, education and general cognition, and smoking were included in the PGS sample. A detailed description of early-life circumstances is presented in eAppendix B.

### Cognitive Outcomes

The HRS assessed cognitive function with a range of tests adapted from the Telephone Interview for Cognitive Status (TICS), which has high validity among non-Hispanic White and non-Hispanic Black adults.^24, 25^ Specifically, the 27-point cognition scale represents three cognitive tests: an immediate and delayed 10-noun free recall test to measure memory (0 to 20 points); a serial sevens subtraction test to measure working memory (0 to 5 points); and a counting backwards test to measure speed of mental processing (0 to 2 points). Built on existing criteria, in each wave of cognitive tests, two categories were derived from the total summary score for cognition (0 to 27 points): normal (12 to 27 points) and cognitively impaired (0 to 11 points).^25, 26^ The latter group included participants ranging from mild cognitive impairment to dementia. Dementia was not separately analyzed due to an insufficient number of dementia patients in the smaller Trauma and PGS samples.^27^ Proxy responses were excluded to ensure the participants had valid measurements of both cognitive score and cognitive impairment.

### Statistical Analysis

To compare the characteristics between White and Black participants, we used Welch t-tests for continuous variables, Mann-Whitney-Wilcoxon tests for ordinal variables, and Chi-square tests for categorical variables.^28–30^ All tests were two-sided with an alpha level of 0.05 for statistical significance.

We used the Blinder-Oaxaca Decomposition (BOD) to attribute racial disparities in cognitive outcomes between White and Black participants to differences in early-life circumstances *versus* differences in the effects of these circumstances.^31^ An illustration of BOD is in eAppendix C. All decompositions were formulated based on Black participants; thus, the contribution of early-life circumstances is characterized as the expected change in cognitive outcomes of Black participants if they had the same mean predictor values as White participants. BOD was implemented based on a standard linear model decomposition for the continuous outcome, i.e., cognitive score, and a nonlinear model decomposition for the dichotomous outcome, i.e., cognitive impairment.^32^

We conducted decompositions at both overall level (i.e., the racial gap in cognitive outcomes explained by all early-life circumstances combined) and the variable level (i.e., the racial gap to which each individual circumstance independently contributed). Age, sex, and marital status were adjusted for in all decomposition analyses.

We separately decomposed cognitive outcomes using three analytic samples (Figure 1). For each analytic sample, we checked similarities of the results using the *earliest* versus *latest* cognitive assessments. All decomposition analyses were carried out using STATA (version 16.1), while all other statistical analyses were conducted using R.

## Results

### Characteristics of the Study Population

Table 1 shows descriptive statistics for cognitive outcomes, early-life circumstances, and covariates of the study population. Black participants had lower cognitive scores and higher proportion of cognitive impairment than White participants in all samples. In the Core sample, the differences in cognitive scores between White and Black participants using the earliest and latest cognitive assessments were 2.7 (95% CI, 2.5-2.9; *P*<.001) points and 1.5 (95% CI, 1.3-1.7; *P*<.001) points, respectively. The gaps in proportion of cognitive impairment between White and Black participants using the earliest and latest cognitive assessments were 15.7 (95% CI, 14.2-17.3; *P*<.001) percentage points (pp) and 10.7 (95% CI, 8.9-12.6; *P*<.001) pp, respectively.

**Table 1.**
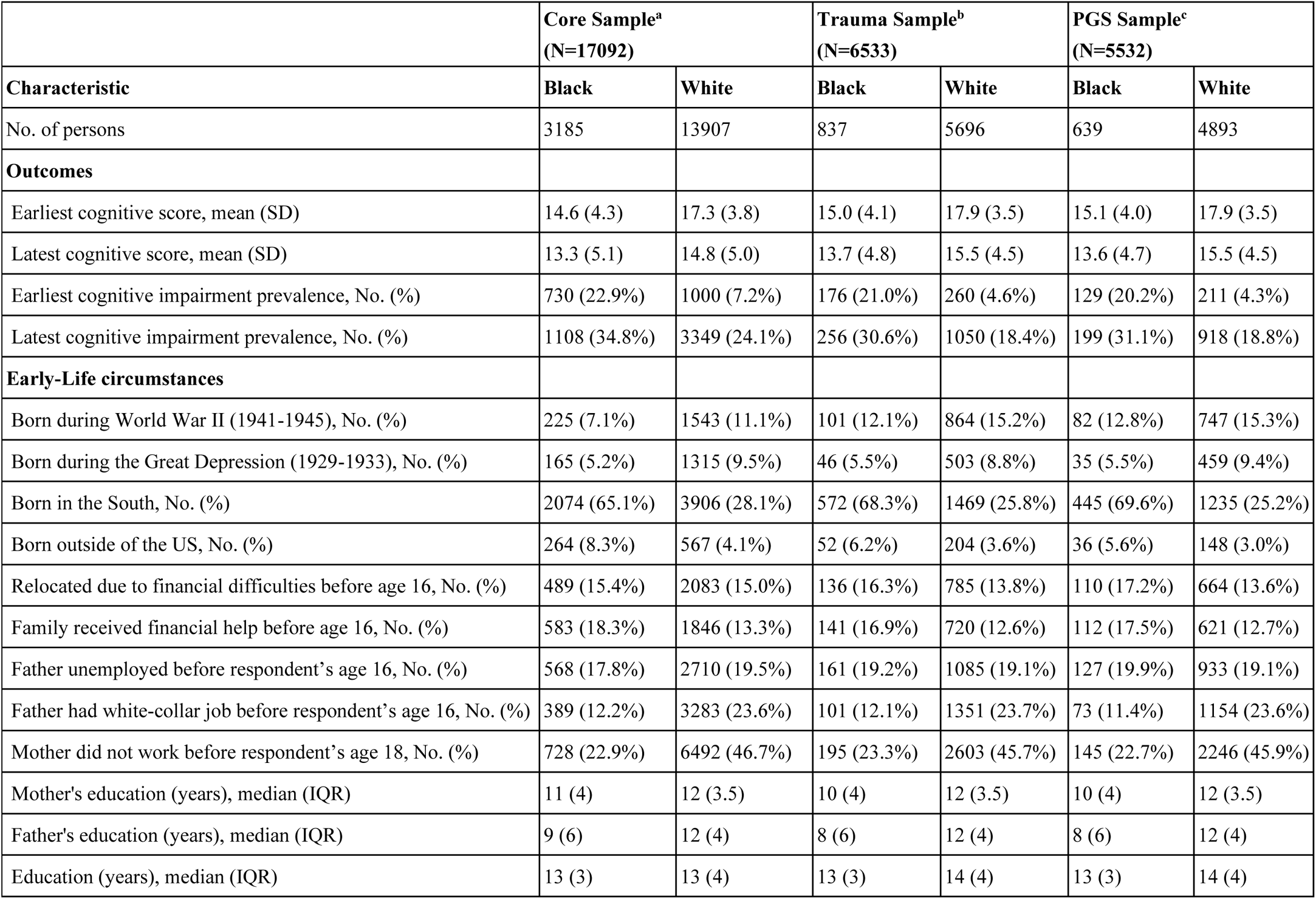
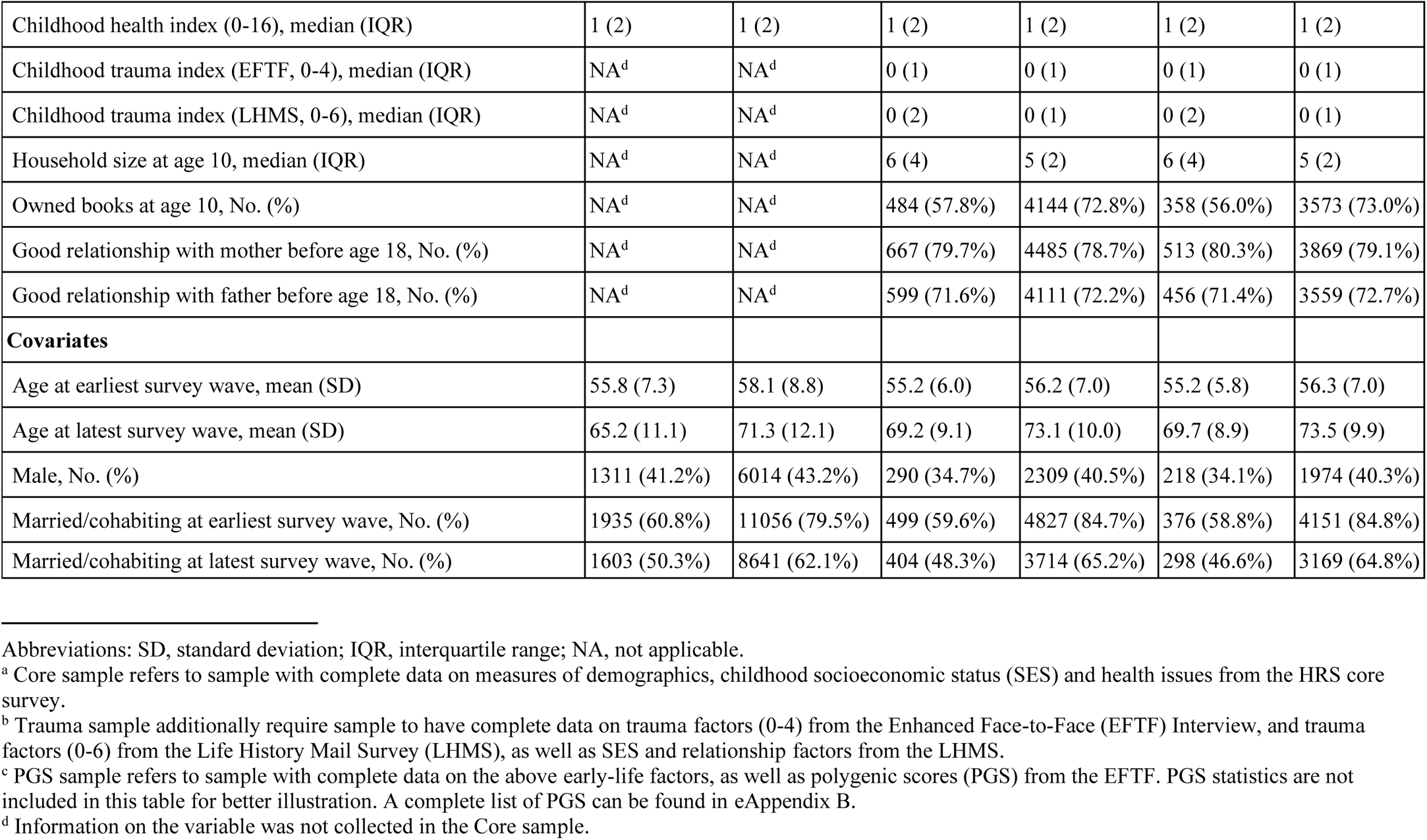
Characteristics of Study Participants

|  | **Core Sample<sup>a</sup>**<br>**(N=17092)** |  | **Trauma Sample<sup>b</sup>**<br>**(N=6533)** |  | **PGS Sample<sup>c</sup>**<br>**(N=5532)** |  |
| --- | --- | --- | --- | --- | --- | --- |
| **Characteristic** | **Black** | **White** | **Black** | **White** | **Black** | **White** |
| No. of persons | 3185 | 13907 | 837 | 5696 | 639 | 4893 |
| **Outcomes** |  |  |  |  |  |  |
| Earliest cognitive score, mean (SD) | 14.6 (4.3) | 17.3 (3.8) | 15.0 (4.1) | 17.9 (3.5) | 15.1 (4.0) | 17.9 (3.5) |
| Latest cognitive score, mean (SD) | 13.3 (5.1) | 14.8 (5.0) | 13.7 (4.8) | 15.5 (4.5) | 13.6 (4.7) | 15.5 (4.5) |
| Earliest cognitive impairment prevalence, No. (%) | 730 (22.9%) | 1000 (7.2%) | 176 (21.0%) | 260 (4.6%) | 129 (20.2%) | 211 (4.3%) |
| Latest cognitive impairment prevalence, No. (%) | 1108 (34.8%) | 3349 (24.1%) | 256 (30.6%) | 1050 (18.4%) | 199 (31.1%) | 918 (18.8%) |
| **Early-Life circumstances** |  |  |  |  |  |  |
| Born during World War II (1941-1945), No. (%) | 225 (7.1%) | 1543 (11.1%) | 101 (12.1%) | 864 (15.2%) | 82 (12.8%) | 747 (15.3%) |
| Born during the Great Depression (1929-1933), No. (%) | 165 (5.2%) | 1315 (9.5%) | 46 (5.5%) | 503 (8.8%) | 35 (5.5%) | 459 (9.4%) |
| Born in the South, No. (%) | 2074 (65.1%) | 3906 (28.1%) | 572 (68.3%) | 1469 (25.8%) | 445 (69.6%) | 1235 (25.2%) |
| Born outside of the US, No. (%) | 264 (8.3%) | 567 (4.1%) | 52 (6.2%) | 204 (3.6%) | 36 (5.6%) | 148 (3.0%) |
| Relocated due to financial difficulties before age 16, No. (%) | 489 (15.4%) | 2083 (15.0%) | 136 (16.3%) | 785 (13.8%) | 110 (17.2%) | 664 (13.6%) |
| Family received financial help before age 16, No. (%) | 583 (18.3%) | 1846 (13.3%) | 141 (16.9%) | 720 (12.6%) | 112 (17.5%) | 621 (12.7%) |
| Father unemployed before respondent's age 16, No. (%) | 568 (17.8%) | 2710 (19.5%) | 161 (19.2%) | 1085 (19.1%) | 127 (19.9%) | 933 (19.1%) |
| Father had white-collar job before respondent's age 16, No. (%) | 389 (12.2%) | 3283 (23.6%) | 101 (12.1%) | 1351 (23.7%) | 73 (11.4%) | 1154 (23.6%) |
| Mother did not work before respondent's age 18, No. (%) | 728 (22.9%) | 6492 (46.7%) | 195 (23.3%) | 2603 (45.7%) | 145 (22.7%) | 2246 (45.9%) |
| Mother's education (years), median (IQR) | 11 (4) | 12 (3.5) | 10 (4) | 12 (3.5) | 10 (4) | 12 (3.5) |
| Father's education (years), median (IQR) | 9 (6) | 12 (4) | 8 (6) | 12 (4) | 8 (6) | 12 (4) |
| Education (years), median (IQR) | 13 (3) | 13 (4) | 13 (3) | 14 (4) | 13 (3) | 14 (4) |
| Childhood health index (0-16), median (IQR) | 1 (2) | 1 (2) | 1 (2) | 1 (2) | 1 (2) | 1 (2) |
| Childhood trauma index (EFTF, 0-4), median (IQR) | NA<sup>d</sup> | NA<sup>d</sup> | 0 (1) | 0 (1) | 0 (1) | 0 (1) |
| Childhood trauma index (LHMS, 0-6), median (IQR) | NA<sup>d</sup> | NA<sup>d</sup> | 0 (2) | 0 (1) | 0 (2) | 0 (1) |
| Household size at age 10, median (IQR) | NA<sup>d</sup> | NA<sup>d</sup> | 6 (4) | 5 (2) | 6 (4) | 5 (2) |
| Owned books at age 10, No. (%) | NA<sup>d</sup> | NA<sup>d</sup> | 484 (57.8%) | 4144 (72.8%) | 358 (56.0%) | 3573 (73.0%) |
| Good relationship with mother before age 18, No. (%) | NA<sup>d</sup> | NA<sup>d</sup> | 667 (79.7%) | 4485 (78.7%) | 513 (80.3%) | 3869 (79.1%) |
| Good relationship with father before age 18, No. (%) | NA<sup>d</sup> | NA<sup>d</sup> | 599 (71.6%) | 4111 (72.2%) | 456 (71.4%) | 3559 (72.7%) |
| **Covariates** |  |  |  |  |  |  |
| Age at earliest survey wave, mean (SD) | 55.8 (7.3) | 58.1 (8.8) | 55.2 (6.0) | 56.2 (7.0) | 55.2 (5.8) | 56.3 (7.0) |
| Age at latest survey wave, mean (SD) | 65.2 (11.1) | 71.3 (12.1) | 69.2 (9.1) | 73.1 (10.0) | 69.7 (8.9) | 73.5 (9.9) |
| Male, No. (%) | 1311 (41.2%) | 6014 (43.2%) | 290 (34.7%) | 2309 (40.5%) | 218 (34.1%) | 1974 (40.3%) |
| Married/cohabiting at earliest survey wave, No. (%) | 1935 (60.8%) | 11056 (79.5%) | 499 (59.6%) | 4827 (84.7%) | 376 (58.8%) | 4151 (84.8%) |
| Married/cohabiting at latest survey wave, No. (%) | 1603 (50.3%) | 8641 (62.1%) | 404 (48.3%) | 3714 (65.2%) | 298 (46.6%) | 3169 (64.8%) |

Abbreviations: SD, standard deviation; IQR, interquartile range; NA, not applicable.
<sup>a</sup> Core sample refers to sample with complete data on measures of demographics, childhood socioeconomic status (SES) and health issues from the HRS core survey.
<sup>b</sup> Trauma sample additionally require sample to have complete data on trauma factors (0-4) from the Enhanced Face-to-Face (EFTF) Interview, and trauma factors (0-6) from the Life History Mail Survey (LHMS), as well as SES and relationship factors from the LHMS.
<sup>c</sup> PGS sample refers to sample with complete data on the above early-life factors, as well as polygenic scores (PGS) from the EFTF. PGS statistics are not included in this table for better illustration. A complete list of PGS can be found in eAppendix B.
<sup>d</sup> Information on the variable was not collected in the Core sample.

Black participants were on average younger than White participants. Their average age gaps in the Core sample using the earliest and latest cognitive assessments, respectively, were 2.4 (95% CI, 2.1-2.7; *P*<.001) and 6.2 (95% CI, 5.7-6.6; *P*<.001) years.

Black participants had less favorable early-life SES than White participants across all samples. In the Core sample, notable differences include a higher chance of receiving financial help from relatives during childhood (Black: 18.3%; White: 13.3%; *P*<.001) and lower levels of maternal education (Black: median=11 years, interquartile range [IQR]=4; White: median=12 years, IQR=3.5; *P*<.001), paternal education (Black: median=9 years, IQR=6; White: median=12 years, IQR=4; *P*<.001) as well as own education (Black: median=13 years, IQR=3; White: median=13 years, IQR=4; *P*<.001). In addition, mothers of Black participants were less likely to not work before respondents’ age of 18 (Black: 22.9%; White: 46.7%; *P*<.001).

Black participants experienced more early-life trauma than White participants. In the Trauma sample, Black participants had a higher score on the 6-point trauma index using early-life factors from the LHMS (Black: median=0 point, IQR=2; White: median=0 point, IQR=1; *P*<.001). Moreover, a lower proportion of Black participants owned books at age 10 (Black: 57.8%; White: 72.8%; *P*<.001); and they had a larger household size at age 10 (Black: median=6 persons, IQR=4; White: median=5 persons, IQR=2; *P*<.001).

### Decomposing Overall Early-life Contributions

The overall contributions of differences in early-life circumstances to disparities between White and Black participants in cognitive score and cognitive impairment are presented in Figure 2 (see eTable 1 for the detailed estimates).

**Figure 2.**
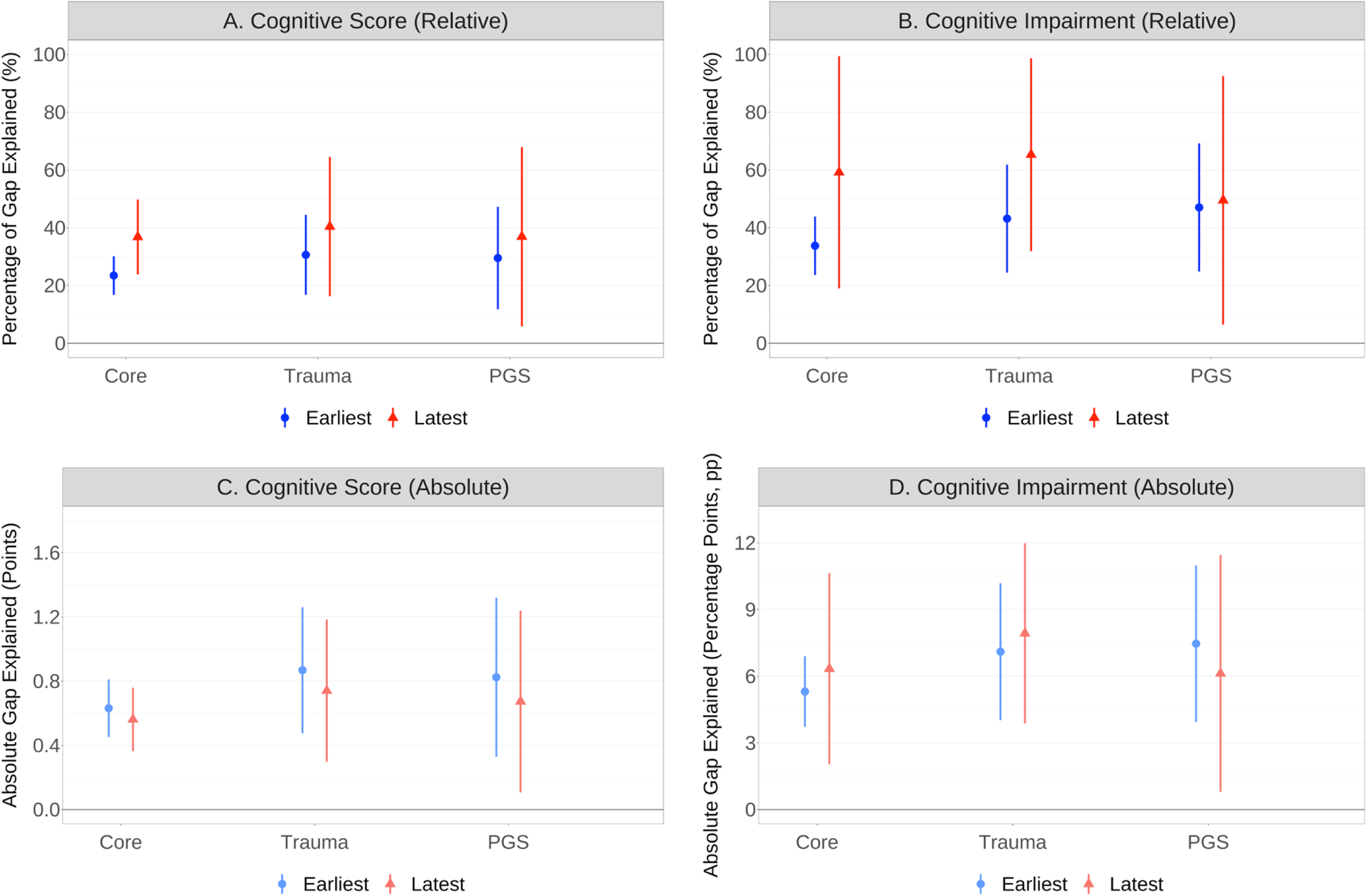
Relative and Absolute Contribution of Overall Early-Life Circumstances to Racial Gaps in Cognition between White and Black Participants. *Notes*: Panel A and C present the decomposition results for cognitive score, a continuous variable ranging from 0 to 27; Panel B and D present the decomposition results for cognitive impairment, a dichotomous variable indicating whether individuals were classified as cognitive impaired (0/1), or not. For each cognitive outcome, X axis denotes the samples being examined, including Core, Trauma, and PGS sample, with an incremental set of early-life circumstances included. In Panel A and B, Y axis denotes the percentage of racial gap in cognition (%) between White and Black participants that was explained by their differences in early-life circumstances, representing the relative contribution of early-life circumstances to racial gaps in cognition. In Panel C and D, Y axis denotes the absolute magnitudes of racial gaps that early-life circumstances explained. For better illustration, the gaps explained in cognitive impairment were presented in absolute values. The point estimates are plotted as circles (*Earliest*) or triangles (*Latest*); and their 95% confidence interval are plotted as vertical lines. For each analytic sample, *Earliest* (in blue color) denotes the sample was analyzed using participants’ earliest cognitive assessments, and *Latest* (in red color) denotes the sample was analyzed using participants’ latest cognitive assessments.

Using the earliest cognitive assessments, differences in early-life circumstances accounted for 0.63 (95% CI, 0.45-0.81) points, or 23.5% of the racial gap in cognitive score between White and Black participants (Figure 2) in the Core sample. The contribution slightly increased as we included more early-life circumstances in the Trauma sample, with differences in early-life circumstances accounting for 0.87 (95% CI, 0.48-1.3) points or 30.6% of the racial gap in cognitive score. In the PGS sample with the richest set of early-life circumstances, differences in early-life circumstances explained 0.82 (95% CI, 0.33-1.3) points or 29.5% of the racial gap in cognitive score. Meanwhile, cognitive impairment exhibited a similar pattern.

Across Core, Trauma, and PGS samples, differences in early-life circumstances explained 5.3 pp (33.8%), 7.1 pp (43.2%), and 7.5 pp (47.0%) of the racial gap in the proportion of cognitive impairment, respectively.

The overall contributions of early-life circumstances were consistently higher when using the latest cognitive assessments than the earliest assessments. Specifically, early-life circumstances across Core, Trauma, and PGS samples explained 0.56 points (36.8%), 0.74 points (40.4%), and 0.67 points (36.9%) of the racial gap in cognitive score, respectively. Moreover. early-life circumstances accounted for 6.3 pp (59.2%), 7.9 pp (65.3%), and 6.1 pp (49.5%) of the racial gap in the proportion of cognitive impairment, respectively.

### Deciphering Main Contributors in Early Life

For the Trauma sample, the early-life circumstances with the greatest individual contributions are presented in Table 2 (absolute) and Figure 3 (relative). Full results for the other samples and variables are provided in eTable 1.

**Figure 3.**
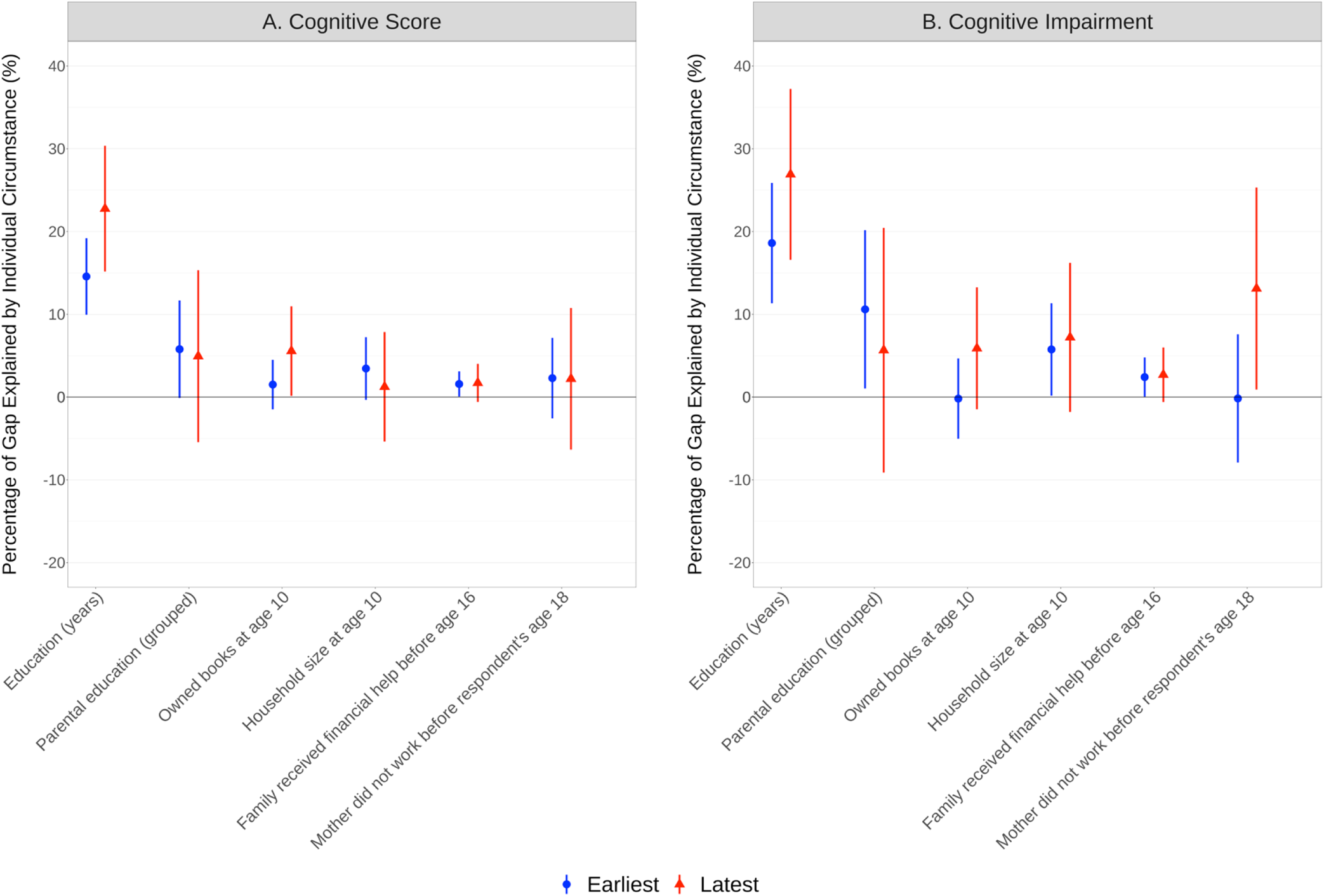
Relative Contribution of Leading Individual Early-Life Circumstances to Racial Gaps in Cognition between White and Black Participants Using the Trauma Sample. *Notes*: Panel A shows the decomposition results for cognitive score, a continuous variable ranging from 0 to 27; Panel B shows the decomposition results for cognitive impairment, a dichotomous variable indicating whether individuals were classified as cognitive impaired (0/1), or not. For each cognitive outcome, X axis denotes the variables being examined; Y axis denotes the percentage of racial gap in cognition (%) between White and Black participants that was explained by their differences in the individual early-life circumstances, representing the relative contribution of individual early-life circumstances to racial gaps in cognition. The relative contributions are plotted as circles (*Earliest*) or triangles (*Latest*); and the 95% confidence interval are plotted as vertical lines. The absolute magnitudes of racial gaps individual early-life circumstances explained are presented in Table 2. For each analytic sample, *Earliest* (in blue color) denotes the sample was analyzed using participants’ earliest cognitive assessments, and *Latest* (in red color) denotes the sample was analyzed using participants’ latest cognitive assessments.

**Table 2.**
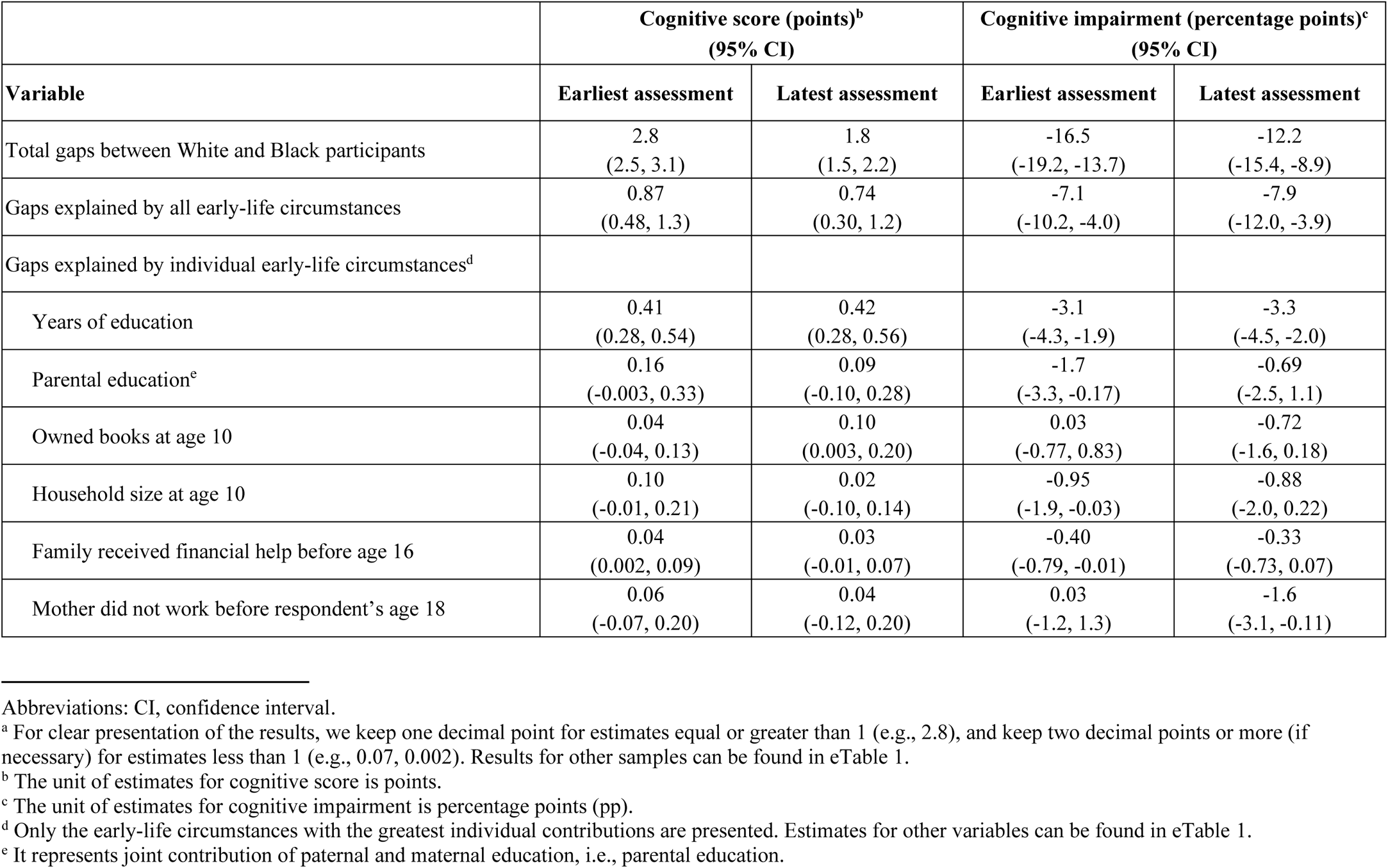
Absolute Contribution of Early-Life Circumstances to Racial Gaps in Cognition Using the Trauma Sample (N=6533)^a^

| Variable | Cognitive score (points)<sup>b</sup><br>(95% CI) |  | Cognitive impairment (percentage points)<sup>c</sup><br>(95% CI) |  |
| --- | --- | --- | --- | --- |
|  | Earliest assessment | Latest assessment | Earliest assessment | Latest assessment |
| Total gaps between White and Black participants | 2.8<br>(2.5, 3.1) | 1.8<br>(1.5, 2.2) | -16.5<br>(-19.2, -13.7) | -12.2<br>(-15.4, -8.9) |
| Gaps explained by all early-life circumstances | 0.87<br>(0.48, 1.3) | 0.74<br>(0.30, 1.2) | -7.1<br>(-10.2, -4.0) | -7.9<br>(-12.0, -3.9) |
| Gaps explained by individual early-life circumstances<sup>d</sup> |  |  |  |  |
| Years of education | 0.41<br>(0.28, 0.54) | 0.42<br>(0.28, 0.56) | -3.1<br>(-4.3, -1.9) | -3.3<br>(-4.5, -2.0) |
| Parental education<sup>e</sup> | 0.16<br>(-0.003, 0.33) | 0.09<br>(-0.10, 0.28) | -1.7<br>(-3.3, -0.17) | -0.69<br>(-2.5, 1.1) |
| Owned books at age 10 | 0.04<br>(-0.04, 0.13) | 0.10<br>(0.003, 0.20) | 0.03<br>(-0.77, 0.83) | -0.72<br>(-1.6, 0.18) |
| Household size at age 10 | 0.10<br>(-0.01, 0.21) | 0.02<br>(-0.10, 0.14) | -0.95<br>(-1.9, -0.03) | -0.88<br>(-2.0, 0.22) |
| Family received financial help before age 16 | 0.04<br>(0.002, 0.09) | 0.03<br>(-0.01, 0.07) | -0.40<br>(-0.79, -0.01) | -0.33<br>(-0.73, 0.07) |
| Mother did not work before respondent's age 18 | 0.06<br>(-0.07, 0.20) | 0.04<br>(-0.12, 0.20) | 0.03<br>(-1.2, 1.3) | -1.6<br>(-3.1, -0.11) |

Abbreviations: CI, confidence interval.
<sup>a</sup> For clear presentation of the results, we keep one decimal point for estimates equal or greater than 1 (e.g., 2.8), and keep two decimal points or more (if necessary) for estimates less than 1 (e.g., 0.07, 0.002). Results for other samples can be found in eTable 1.
<sup>b</sup> The unit of estimates for cognitive score is points.
<sup>c</sup> The unit of estimates for cognitive impairment is percentage points (pp).
<sup>d</sup> Only the early-life circumstances with the greatest individual contributions are presented. Estimates for other variables can be found in eTable 1.
<sup>e</sup> It represents joint contribution of paternal and maternal education, i.e., parental education.

Years of education contributed the most to racial gaps in cognitive outcomes. Using the earliest cognitive assessments, years of education explained 0.41 points or 14.6% of the racial gap in cognitive score, and 3.1 pp or 18.6% of the racial gap in the proportion of cognitive impairment. The contribution of years of education increased when using the latest cognitive assessments, accounting for 0.42 points or 22.8% of the racial gap in cognitive score, and 3.3 pp or 26.9% of the racial gap in the proportion of cognitive impairment.

Early-life educational environments also greatly contributed to the racial disparities. Using the latest cognitive assessments, owning books at age 10 explained 0.10 points or 5.6% of the racial gap in cognitive score, while having a mother who did not work accounted for 1.6 pp or 13.1% of the racial gap in the proportion of cognitive impairment. Parental education jointly explained 1.7 pp or 10.6% of the racial gap in cognitive impairment when using the earliest cognitive assessments.

Family receiving financial help explained 0.04 points or 1.6% of the racial gap in cognitive score and 0.40 pp or 2.4% of the racial gap in proportion of cognitive impairment, when using the earliest cognitive assessments. The effect of household size at age 10 was small and inconsistently observed for the two cognitive outcomes.

Finally, the place or timing of the birth, childhood adverse health events, and childhood trauma were not significant contributors to racial gaps in cognition in the Trauma sample, though some factors, such as being born in the South and childhood adverse health events, were associated with racial gaps in the Core sample (see eTable 1). Polygenic scores for cognitive outcomes only had small and insignificant contributions when included in the PGS sample (eTable 1).

## Discussion

In this prospective longitudinal study of American older adults, we evaluated to what extent two clinically meaningful cognitive outcomes differ between Black and White participants, and quantified how much early-life circumstances contributed collectively and individually to the racial gaps. Four major findings warrant comment.

First, the less favorable early-life social-environmental circumstances among Black participants relative to White participants explained a substantial portion of their gaps in cognitive outcomes in the US.^33–36^ Moreover, by inclusion of the polygenic scores, biological susceptibilities were considered in this analysis.^37–40^ Our findings suggest that addressing intergenerational transmission of structural inequalities could improve cognitive well-being of older persons and its racial disparities.

Second, among all early-life circumstances investigated, lower educational attainment and factors of disadvantaged family educational environment among Black participants (including owning no books at home, low parental education, having a working mother) were the greatest contributors to racial gaps in cognitive outcomes, even after considering a rich set of additional risk factors, including early-life SES, demographics, health, trauma, and genetic factors. Educational attainment and environment may work through a comprehensive set of channels to influence racial gaps, such as brain development, employment, income, access to health care, and lifestyle choices. For example, non-working mothers may possibly have spent more time with their children, contributing to their early brain development.^41^ Overall, our results suggest that policies implemented to improve equitable childhood education may generate long-lasting impacts on reducing racial disparities in cognitive outcomes into older ages.

While the 2020 LANCET Commission Report ranked educational attainment among the most important and potentially modifiable risk factors in adulthood for cognitive impairment,^42^ few prior studies have evaluated the associations between a comprehensive set of early-life circumstances, including factors of early educational environment, and racial gaps in cognitive outcomes.^3, 43–45^

Third, demographic and SES factors, including household size (indicating potential competition for resources) and financial difficulty, were additional contributors to the racial gap in cognitive outcomes.^33, 34, 36, 46^ While place of birth (U.S. South) was not found to be a significant contributor to racial disparities in our preferred Trauma sample, future work should use finer geographic data to dissect essential mechanisms attached to the places that may shape the racial gap in cognition.^33, 34, 36, 47–49^ Overall, these results suggest that promoting more equitable social and economic opportunities are required to address long-term racial inequities.^33, 34^

Fourth, the relative contributions of overall and leading early-life circumstances to racial gaps in cognitive outcomes among survivors tended to increase over time, which further emphasizes the importance of intervening earlier in life to mitigate growing gaps over the life span.

It is also worth noting that the rising socioeconomic disparities in recent decades were not considered in the current analysis, which focused on a relatively older cohort.^33, 34, 36, 50^ Greater socioeconomic disparities also tend to transmit disadvantages from parents to children to affect long-term health among the latter.^51^ Consequently, the large racial gaps in cognitive outcomes may worsen despite safety net programs being in place.^52^ It is therefore important to monitor the trend in racial gaps for future age cohorts and generations.

This analysis builds on the existing evidence on education and later-life cognitive outcomes, especially studies using the same validated TICS in HRS or exploring racial/ethnic differences in the education-cognition linkage.^53–56^ An important strength of the current analysis is our advancement of the literature on the role of early-life circumstances prior to full educational attainment. A second main strength relates to our accounting for a large number of early-life circumstances, which enables us to better understand their independent role and how they were coalesced to shape racial disparities in late-life cognition. To our knowledge, this represents the first quantification of the contributions made by this uniquely rich set of individual-level early-life risk factors on black-white disparities in cognitive outcomes in old age. The third main strength of this study involves long durations of follow-up that offer us novel opportunities to assess the evolving role of early-life circumstances on racial gaps in cognition over time.

### Limitations

Our observational study has several limitations. First, persons with impaired cognition may have been less likely to participate in the HRS baseline assessment or to complete follow-up assessments because of attrition or death. Mortality selection may bias against including respondents with more disadvantaged early-life circumstances, suggesting that the associations between early-life circumstances and late-life racial disparities in cognition may be conservative. Second, early-life circumstances in this study were self-reported, which may introduce recall bias. Third, due to the small sample size for other racial/ethnic groups, the current study focused solely on comparisons between Black and White participants. The low prevalence of dementia prevented us from investigating its early-life contributors. Fourth, our model may suffer from overfitting due to the inclusion of a large set of early-life circumstances. Fifth, using increasingly granular information from the Core to the PGS samples, these models were not adjusted for post-hoc multiple comparisons. Finally, while there is temporality in exposures and subsequent cognitive outcomes, the associations should not be interpreted as causal. Future work may identify causal channels to inform policies and interventions, for example, if the effects are directly driven by early-life environments or through parental transmission of unhealthy lifestyles and other choices.

## Conclusions

This study identified early-life circumstances that may contribute to racial disparities in cognitive outcomes among American older adults. Future analyses are required to elucidate the mechanisms and inform the development of early-life interventions to slow the process of cognitive impairment and help address its disparities across racial groups.

## Data Availability

All data produced are available online at
https://hrs.isr.umich.edu/

## Acknowledgements

This work was supported by the Career Development Award (K01AG053408) from the National Institute on Aging; Claude D. Pepper Older Americans Independence Center at Yale School of Medicine, funded by the National Institute on Aging (P30AG021342); and Yale Alzheimer’s Disease Research Center (P30AG066508); James Tobin Research Fund at Yale Economics Department; and Yale Macmillan Center Faculty Research Award. The funders had no role in the study design; data collection, analysis, or interpretation; in the writing of the report; or in the decision to submit the article for publication. The authors acknowledge helpful comments by participants and discussants at the various conferences, seminars, and workshops.

## Supplementary Online Content

### eAppendix A. Data Source

#### Data source 1. The HRS Core Survey

The HRS study has biennial interviews for collecting a wide range of information, including economics, health, marital, family status, and public and private support systems from 1992 to 2018 (the most recent wave).^1^ Although the HRS has grown with the addition of new cohorts, the contents of the core survey have remained mostly consistent. In particular, the HRS core survey generally included multiple sections such as demographics and background, health, cognition, family structure and transfers, functional limitations, housing, physical measures, employment and pensions, disability, health services and insurance, expectations, assets and income, assets change, widowhood and divorce, and insurance. Additionally, the HRS has some experimental modules on specialized topics as part of the core survey. These modules only target a random subsample at the end of the core survey.^1, 2^

In this study, variables of early-life socioeconomic status (SES) were constructed using the measures from the HRS core survey and/or related modules. Many of the SES variables were assembled from the RAND HRS files since RAND HRS groups have created user-friendly files with cleaned and processed variables with consistent and intuitive naming conventions, as well as model-based imputations.^3^ Otherwise, we assembled variables from the original HRS released core data from 1996 (when the variables were available) to 2018.

#### Data source 2. The Life History Mail Survey

The HRS Life History Mail Survey (LHMS) contains additional questions about respondents’ residential history, educational history, and other important early-life and family events. The 2015, 2017 Spring, and 2017 Fall versions were conducted in subsamples of HRS participants (2015: n=11256; 2017 Spring: n=5174; 2017 Fall: n=5180). The target subsample for the 2015 wave included all living HRS participants who were not included in the 2015 Consumption and Activities Mail Survey (CAMS) and who completed their most recent HRS core survey interview in English (rather than Spanish). The 2017 Spring wave included participants who were 2015 CAMS Sample Members who were still alive in 2017 and whose household was considered finalized on their 2016 core interview(s) by early March 2017 (members of finalized households either had completed core interviews or were considered final refusals for the core that wave). Lastly, the 2017 Fall sample included participants who were not included in the 2017 CAMS sample, and who did not return a 2015 LHMS questionnaire. The response rates and total enrolled participants for the 2015, 2017 Spring, and 2017 Fall waves respectively were 58% and 6,481 participants, 74% and 3,844 participants, and 28% and 1,444 participants.^4–6^ We assembled many variables for early-life traumas and health behaviors contained in all three waves of the LHMS.

#### Data source 3. The Enhanced Face-to-Face Interview

In 2006, HRS initiated the Enhanced Face-to-Face (EFTF) interview with a mixed-mode design during the follow-up period in which a random half of HRS participants were assigned a face-to-face interview with physical and biological measures (e.g., salivary DNA samples) and a psychosocial questionnaire. The other half completed only the Core survey but were selected for the next (i.e., 2008) EFTF interview. A similar method was applied to the subsequent HRS survey. In fact, the psychosocial questionnaire was piloted in 2004 and included personal evaluations of their life circumstances, subjective well-being, lifestyle and stress. Participants completed it and returned by mail. Several early-life circumstance variables in the present study related to trauma were assembled from this psychosocial questionnaire from 2006 to 2016.

In addition, polygenic scores for a variety of phenotypes from respondents who provided salivary DNA between 2006 and 2012 were included in this study. Genotyping was conducted by the Center for Inherited Disease Research (CIDR) in 2011, 2012, and 2015, and principal component analysis was performed to identify population group outliers and to provide sample eigenvectors for association testing to adjust for potential population stratification. The final European American sample (n=12,090) included all participants who self-reported as non-Hispanic White that had PC loadings within ± one standard deviations of the mean for eigenvectors 1 and 2 in the PC analysis of all unrelated study subjects. The final African American sample (n=3,100) included all self-reported non-Hispanic African Americans within two standard deviations of the mean of all self-identified African Americans for eigenvector 1 and ± one standard deviation of the mean for eigenvector 2 in the PC analysis of all unrelated study subjects.^7^

### eAppendix B. Early-Life Circumstances

Early-life circumstances involve all exposures during the early stages of life that may matter to long-term health outcomes. This study classified all early-life circumstances into the domains of demographics, SES, family relationships, trauma, health, and genetics. Domains were defined as a group of exposures. Some definitions are described below.

#### Domain 1. Early-life Socioeconomic status (SES)

We included several relevant variables that have been used before,^8, 9^ such as whether the respondent relocated due to financial difficulties, whether their family had received financial assistance, and the respondents’ fathers’ and mothers’ highest level of education in years. One additional variable—“father lost job”, indicating that the participants’ fathers experienced a significant unemployment spell (“several months or more”)—was included, as it was suggested to appropriately reflect early-life SES.^10^ Moreover, to incorporate information on parental occupation, whether the respondent’s father worked a white-collar job and whether the respondent’s mother was employed during the respondent’s childhood were also included. Finally, we additionally incorporated a variable— “owned books at age 10” —from the LHMS, which further provided insight into respondents’ early life educational environments.^11–13^

#### Domain 2. Early-life Traumas

We assembled several indicators of childhood traumatized experiences from the EFTF and the LHMS, which have been used before.^14^ Each variable for childhood trauma was coded as a dichotomous variable based on the answer (0=no; 1=yes). To obtain a summarized score, we summed them in an unweighted way as a total score. Since these trauma variables were from two different data sources (i.e., the EFTF interview as well as the 2015 Fall, 2017 Spring, and 2017 Fall LHMS), there was a slight difference in age frame in the questions (18 years old vs. age 16); hence we constructed two trauma indices consisting of measures from each respective source. The trauma index consisting of measures from the EFTF was scored from 0-4, while the one with measures from the LHMS was scored from 0-6. Higher scores represent more trauma burden. The individual traumas included in our study from each data source are given below:

### Traumas included from the EFTF

- Physical abused prior to age 18
- Parents used drugs or alcohol which caused problems prior to age 18
- Trouble with police prior to age 18
- Repeated school prior to age 18

### Traumas included from the LHMS

- Spent any amount of time in an orphanage prior to age 16
- Spent any amount of time in a foster home prior to age 16
- One or more parents died prior to age 16
- Parents divorced prior to age 16
- Separated from mother for more than 6 months prior to age 16
- Separated from father for more than 6 months prior to age 16

#### Domain 3. Early-life Health Events

To assess early-life health, we included 16 important health events occurring prior to age 18 from the Core, such as disabled for six months and severe head injury.^9, 15^ Each variable for childhood health was coded as a dichotomous variable (0=no; 1=yes), and to obtain a summarized measure, we used an unweighted sum (possible range 0-16). All health outcomes which we included are as follows:

- Disabled for six months or more prior to age 18
- Injury to head which resulted in the loss of consciousness prior to age 18
- High blood pressure prior to age 18
- Heart issues prior to age 18
- Respiratory issues prior to age 18
- Diabetes prior to age 18
- Epilepsy prior to age 18
- Problems with speech prior to age 18
- Problems with hearing prior to age 18
- Problems with vision prior to age 18
- Depressed prior to age 18
- Other emotional or psychological disorder prior to age 18
- Learning disability prior to age 18
- Usage of drugs or alcohol prior to age 18
- Smoking cigarettes prior to age 18
- Parents smoked cigarettes prior to age 18

#### Missing Responses

To reduce sample attrition, missing responses in the domains of severe early-life traumas and health issues were re-coded to zero for a small number of participants who did not respond to those questions in our main analysis. While these traumas and health issues are rare, a sensitivity analysis was conducted using samples without re-coding to account for the potential bias and the estimates were consistent with our main findings.

#### Domain 4. Genetic factors

Polygenic score (PGS), also known as polygenic risk score or genetic risk score, is a value based on variation in multiple genetic loci and their associated weights. It serves as the best prediction for the trait that can be made when accounting for variation in multiple genetic variants. All phenotypes for the five domains for genetic factors are listed below, and more information can be found in the document from HRS.^7^ We controlled for potential population stratification.^16^

| **Polygenic score (PGS)** | **Traits/phenotypes** |
| --- | --- |
| Anthropometric | Height |
|  | Body mass index |
|  | Waist circumference |
|  | Waist-to-Hip ratio |
| Disease/longevity | Coronary artery disease |
|  | Myocardial infarction |
|  | Type II diabetes |
|  | Alzheimer's disease IGAP 2013 with APOE |
|  | Alzheimer's disease IGAP 2019 with APOE |
|  | Longevity |
| Mental health/personality | Attention deficit/hyperactivity disorder PGC 2010 |
|  | Attention deficit/hyperactivity disorder PGC 2017 |
|  | Autism |
|  | Bipolar disorder |
|  | Mental health cross disorder |
|  | Major depressive disorder PGC 2010 |
|  | Major depressive disorder PGC 2018 |
|  | Subjective wellbeing |
|  | Neuroticism |
|  | Depressive symptoms |
|  | Schizophrenia |
|  | Extraversion |
| Education/cognition | Educational attainment |
|  | General cognition 2015 |
|  | General cognition 2018 |
| Smoking | Smoking initiation (ever/never) |
|  | Number of cigarettes smoked per day TAG 2010 |
|  | Number of cigarettes smoked per day GSCAN 2019 |

#### Domains 5&6. Demographics & Family Relationships

Demographic factors included birth in and outside of the U.S., birth in the U.S. South, birth during the Great Depression (1929-1933), birth during World War II (1941-1945), and household size at age 10. Familial relationships included respondents’ self-rated quality of their relationship with their parents during childhood. Specifically, the relationship measures were coded as dichotomous variables (0=no; 1=yes) to indicate whether the respondent had good relationship with parents or not before age 18.

### eAppendix C. Decomposition Analysis

#### Blinder-Oaxaca Decomposition (BOD)

was used to measure the extent to which early-life circumstances may individually and collectively contribute to racial disparities in cognitive outcomes, including cognitive score and cognitive impairment, between *White* and *Black* participants. The BOD decomposes mean differences in regression outcomes in a counterfactual manner, and is widely used in economics to understand racial disparities.^17–27^ It allows us to separate cognitive differences between White and Black participants into a part that is explained by differences in early-life circumstances, versus a part that cannot be accounted for by such differences.^19, 28^ We tested the hypothesis that early-life circumstances contribute significantly and sizably to racial disparities in cognitive outcomes.

We focused on comparing racial disparities in cognitive outcomes between White and Black participants, and examined the extent to which racial differences are attributed to the differences in early-life circumstances.^29, 30^ Formally, the differences in cognitive outcomes between White (*W*) and Black (*B*) participants can be formulated as,

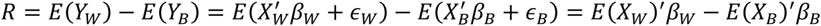

where *E*(*Y*_*l*_) denotes the expected value of cognitive outcomes for race *l*, *l* ∈ (*W*, *B*); and *X*_*l*_ is a matrix that contains early life circumstances and constants. The second and the third equalities hold under the assumptions of linear model, 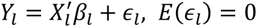. The equation can be decomposed as follows.

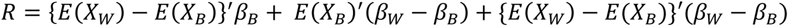

The first component {*E*(*X*_*W*_) − *E*(*X*_*B*_)}^′^*β*_*B*_ represents the part of the differential that is attributable to group differences in early-life circumstances. The second component *E*(*X*_*B*_)^′^(*β*_*W*_ − *β*_*B*_) measures the contribution of differences in coefficients. The third component {*E*(*X*_*W*_) − *E*(*X*_*B*_)}′(*β*_*W*_ − *β*_*B*_) is an interaction term accounting for both the differences in early-life circumstances and coefficients. Our parameter of interest is the first component, which measures the expected changes in Black participants’ mean cognition if they had the same levels of early life circumstances as White participants (i.e., overall contribution of early-life circumstances). To understand how much of the racial disparities in cognitive outcomes are accounted for by individual characteristics, this component can be further decomposed as,

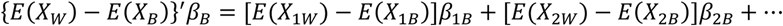

where *E*(*X*_1_), *E*(*X*_2_), … are the expectation of individual characteristics; and *β*_1_, *β*_2_, … are the associated coefficients. [*E*(*X*_1*W*_) − *E*(*X*_1*B*_)]*β*_1*B*_, thus captures the individual contribution of the differences in *X*_1_.

In this study, the decompositions were conducted at both the overall and the variable level to evaluate the collective and individual contribution of early-life circumstances. For continuous outcome (i.e., cognitive score), BOD was conducted using a linear decomposition method, while for dichotomous outcomes (i.e., cognitive impairment, dementia), a nonlinear decomposition method was employed.^17–27, 31^

---

**eFigure 1.**
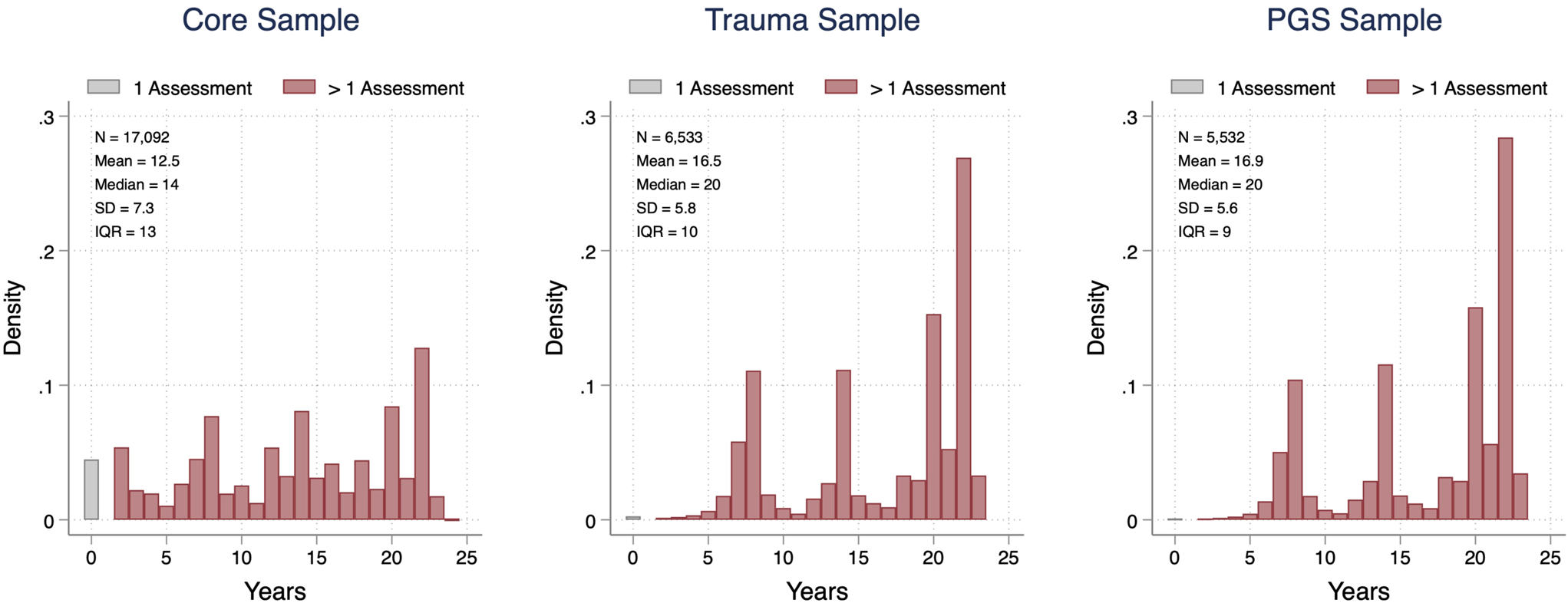
Years Gap between the Earliest and the Latest Cognitive Assessment. *Notes*: “1 Assessment” refers to the participants with only one cognitive assessment between 1995-2018; “>1 Assessment” refers to the participants with more than 1 cognitive assessment between 1995-2018. Abbreviations: SD, standard deviation; IQR, interquartile range.

**eTable 1a.**
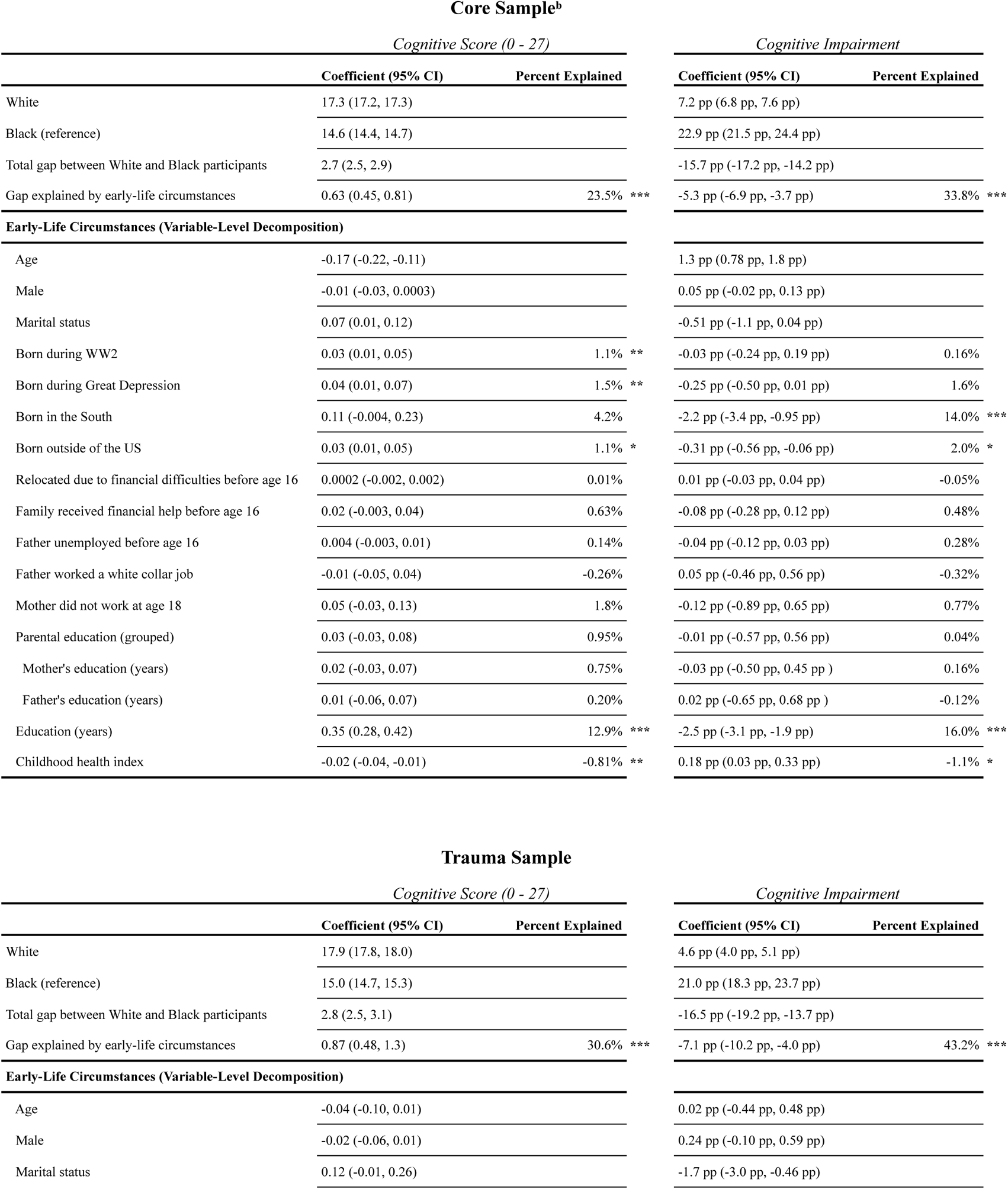
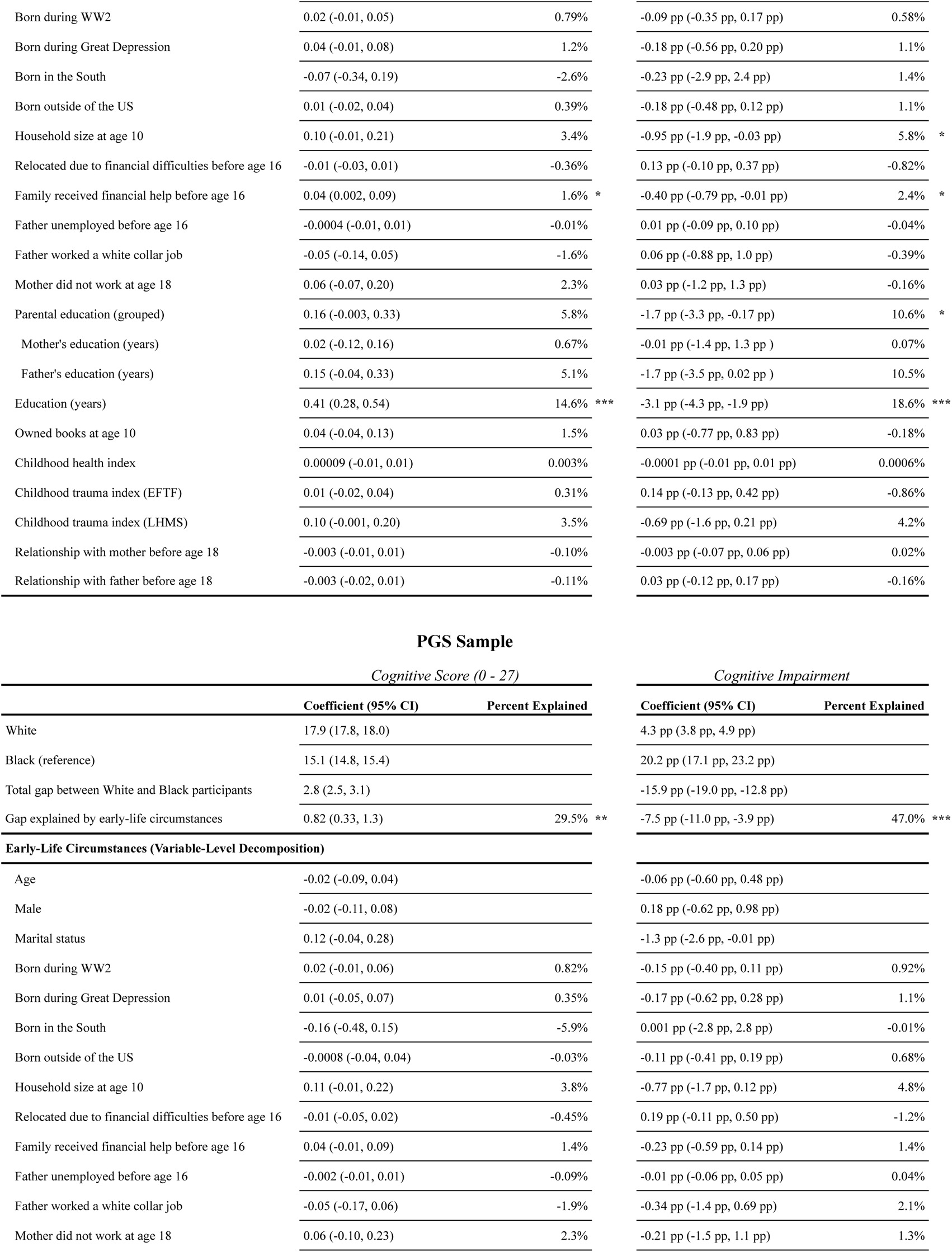
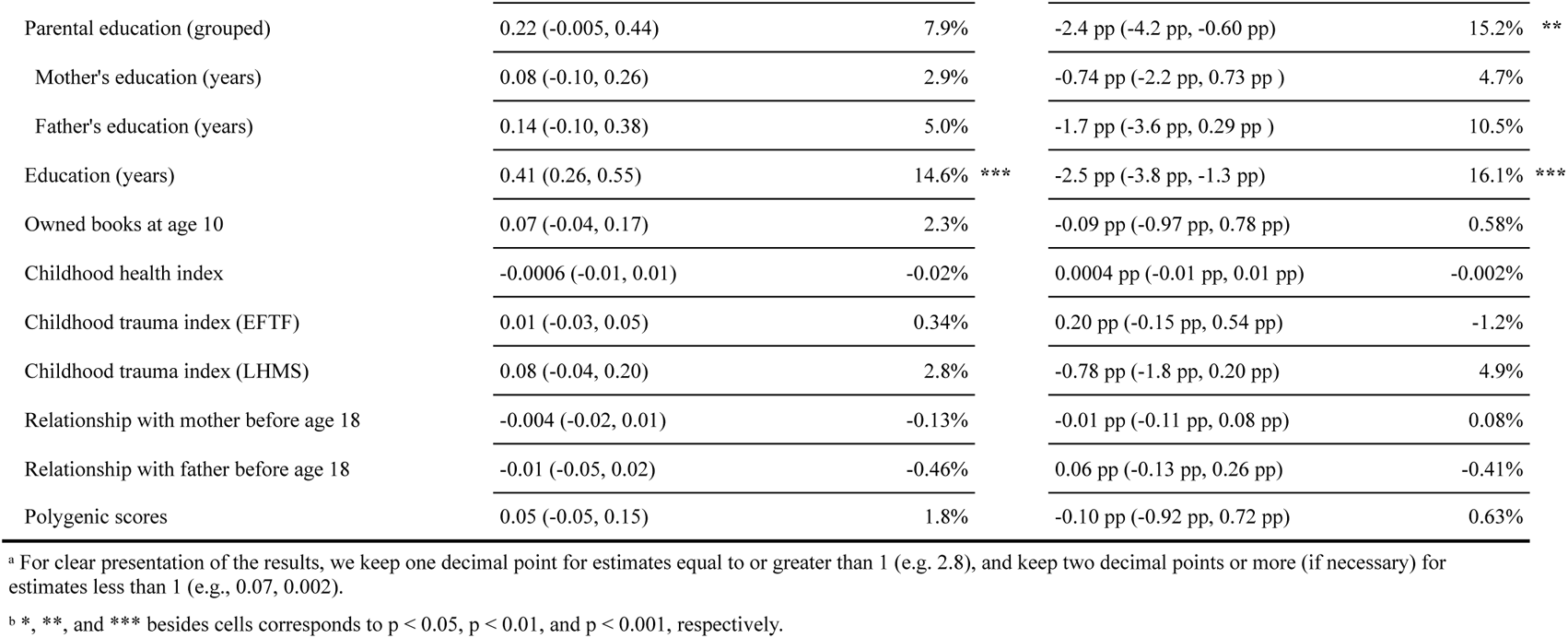
Decomposition Results Using Respondents’ Earliest Recorded Data^a^

**eTable 1b.**
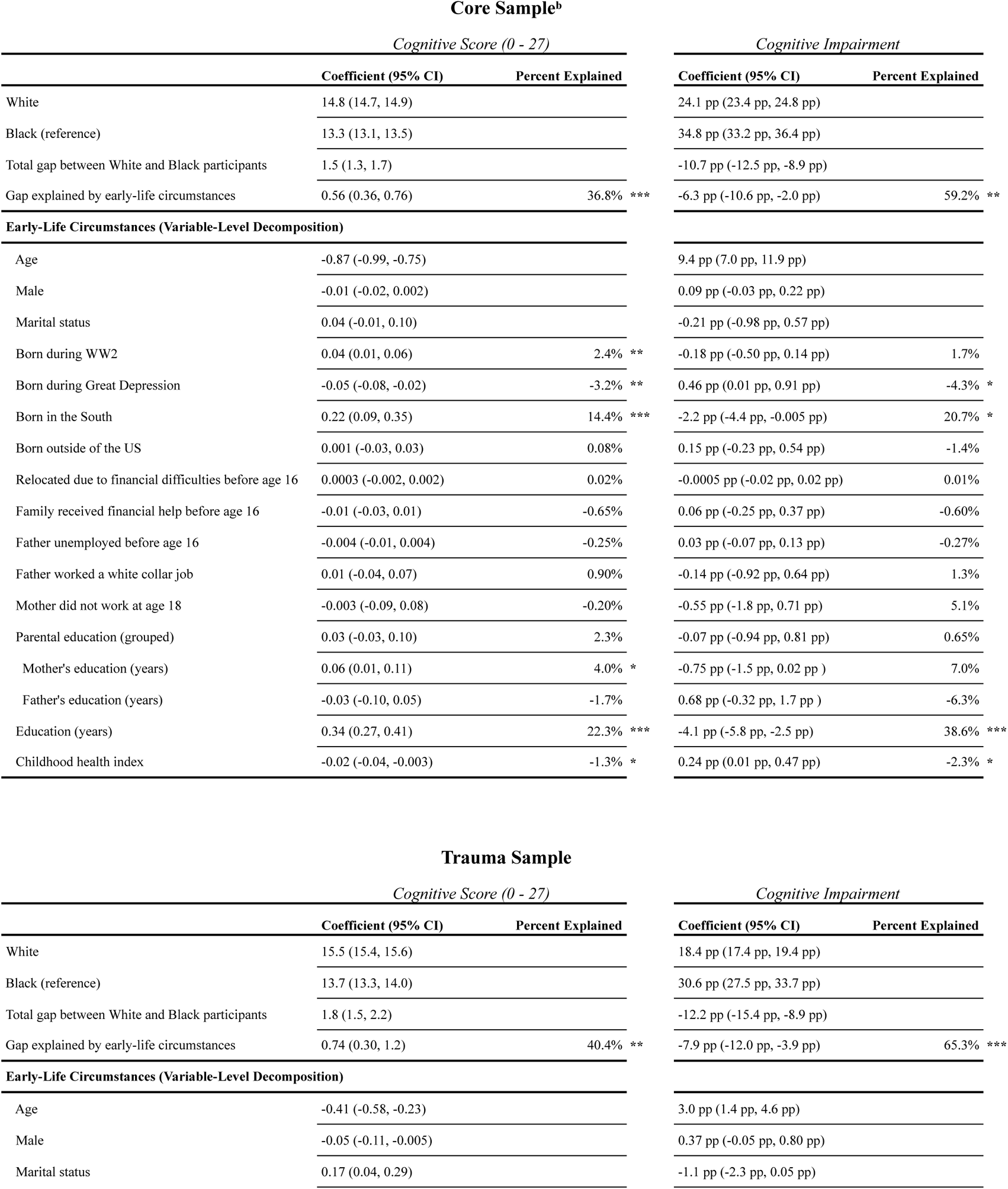
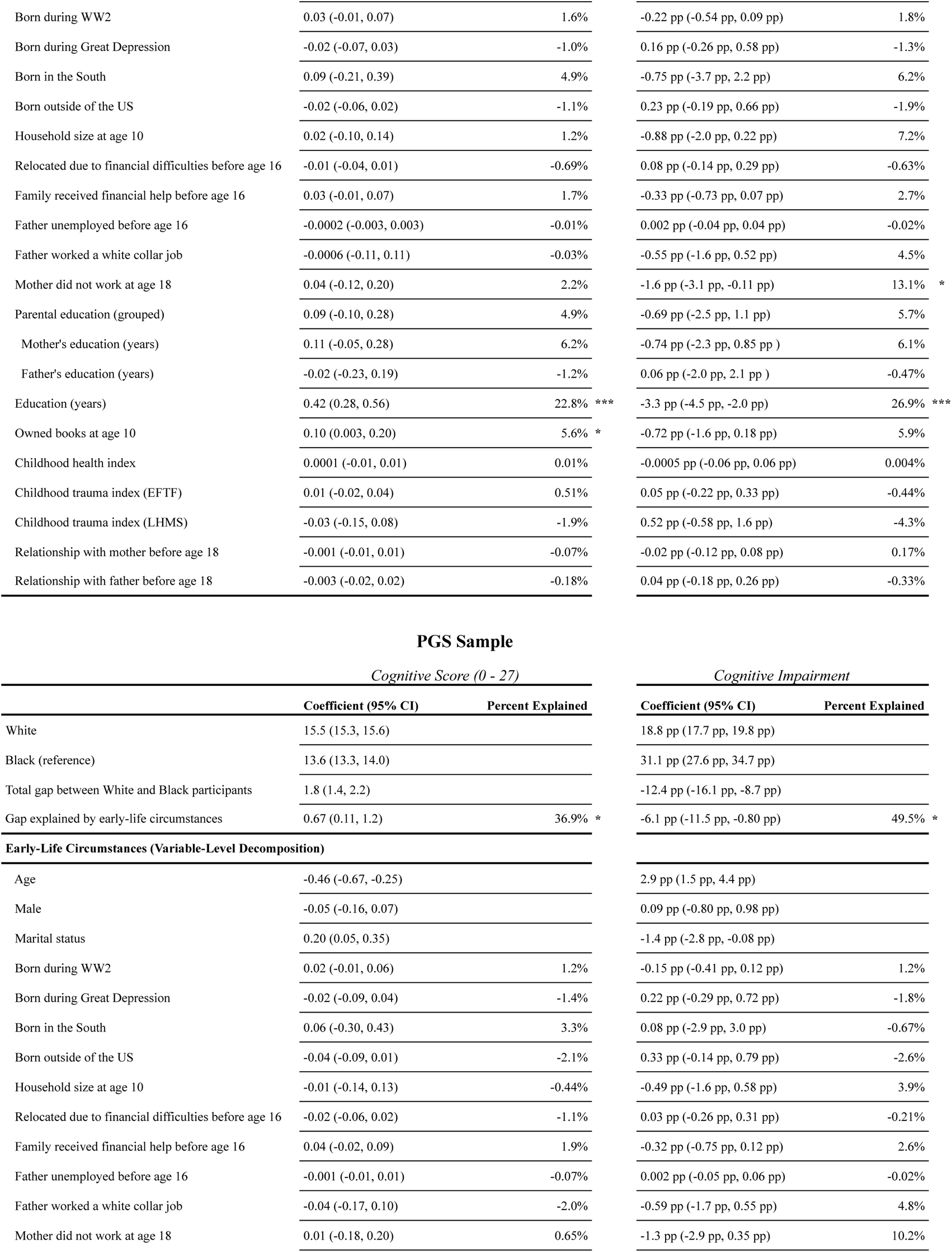
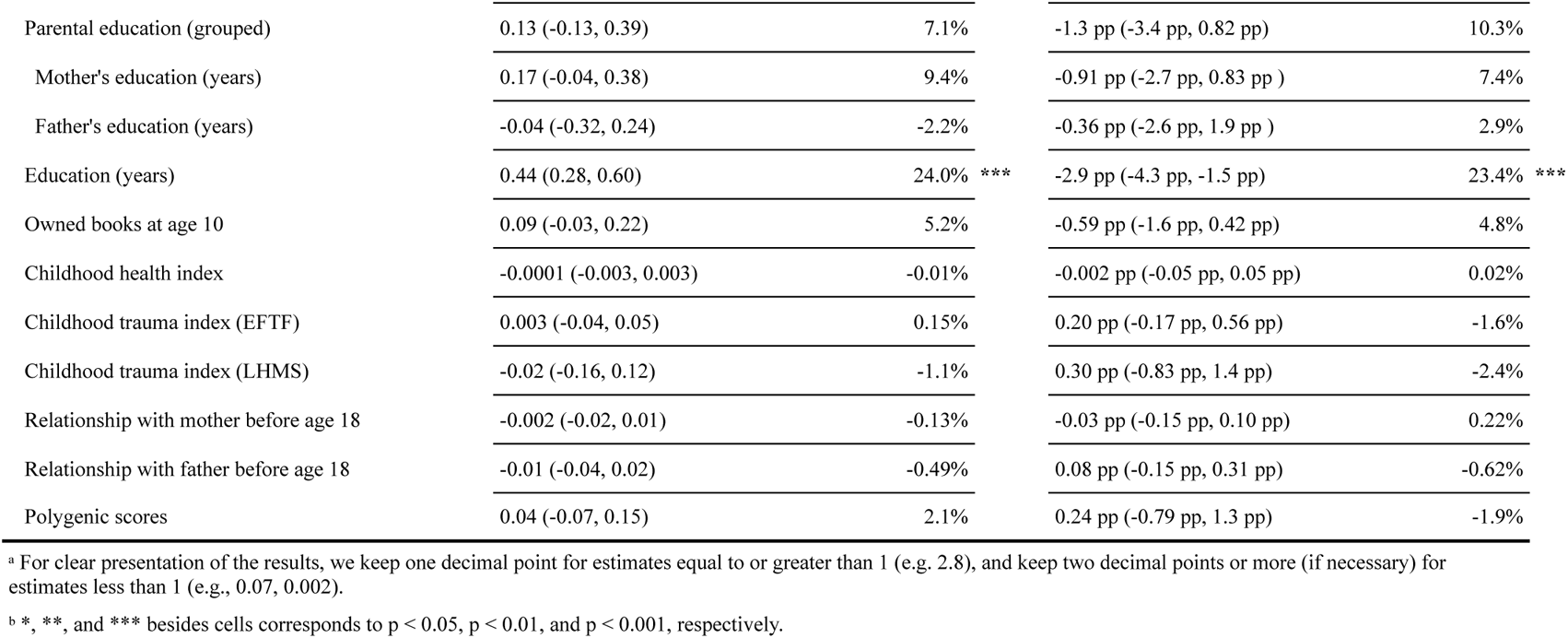
Decomposition Results Using Respondents’ Latest Recorded Data^a^

| Core Sample<sup>b</sup> |  |  |  |  |
| --- | --- | --- | --- | --- |
| Cognitive Score (0 - 27) |  |  | Cognitive Impairment |  |
|  | Coefficient (95% CI) | Percent Explained | Coefficient (95% CI) | Percent Explained |
| White | 14.8 (14.7, 14.9) |  | 24.1 pp (23.4 pp, 24.8 pp) |  |
| Black (reference) | 13.3 (13.1, 13.5) |  | 34.8 pp (33.2 pp, 36.4 pp) |  |
| Total gap between White and Black participants | 1.5 (1.3, 1.7) |  | -10.7 pp (-12.5 pp, -8.9 pp) |  |
| Gap explained by early-life circumstances | 0.56 (0.36, 0.76) | 36.8% *** | -6.3 pp (-10.6 pp, -2.0 pp) | 59.2% ** |
| Early-Life Circumstances (Variable-Level Decomposition) |  |  |  |  |
| Age | -0.87 (-0.99, -0.75) |  | 9.4 pp (7.0 pp, 11.9 pp) |  |
| Male | -0.01 (-0.02, 0.002) |  | 0.09 pp (-0.03 pp, 0.22 pp) |  |
| Marital status | 0.04 (-0.01, 0.10) |  | -0.21 pp (-0.98 pp, 0.57 pp) |  |
| Born during WW2 | 0.04 (0.01, 0.06) | 2.4% ** | -0.18 pp (-0.50 pp, 0.14 pp) | 1.7% |
| Born during Great Depression | -0.05 (-0.08, -0.02) | -3.2% ** | 0.46 pp (0.01 pp, 0.91 pp) | -4.3% * |
| Born in the South | 0.22 (0.09, 0.35) | 14.4% *** | -2.2 pp (-4.4 pp, -0.005 pp) | 20.7% * |
| Born outside of the US | 0.001 (-0.03, 0.03) | 0.08% | 0.15 pp (-0.23 pp, 0.54 pp) | -1.4% |
| Relocated due to financial difficulties before age 16 | 0.0003 (-0.002, 0.002) | 0.02% | -0.0005 pp (-0.02 pp, 0.02 pp) | 0.01% |
| Family received financial help before age 16 | -0.01 (-0.03, 0.01) | -0.65% | 0.06 pp (-0.25 pp, 0.37 pp) | -0.60% |
| Father unemployed before age 16 | -0.004 (-0.01, 0.004) | -0.25% | 0.03 pp (-0.07 pp, 0.13 pp) | -0.27% |
| Father worked a white collar job | 0.01 (-0.04, 0.07) | 0.90% | -0.14 pp (-0.92 pp, 0.64 pp) | 1.3% |
| Mother did not work at age 18 | -0.003 (-0.09, 0.08) | -0.20% | -0.55 pp (-1.8 pp, 0.71 pp) | 5.1% |
| Parental education (grouped) | 0.03 (-0.03, 0.10) | 2.3% | -0.07 pp (-0.94 pp, 0.81 pp) | 0.65% |
| Mother's education (years) | 0.06 (0.01, 0.11) | 4.0% * | -0.75 pp (-1.5 pp, 0.02 pp ) | 7.0% |
| Father's education (years) | -0.03 (-0.10, 0.05) | -1.7% | 0.68 pp (-0.32 pp, 1.7 pp ) | -6.3% |
| Education (years) | 0.34 (0.27, 0.41) | 22.3% *** | -4.1 pp (-5.8 pp, -2.5 pp) | 38.6% *** |
| Childhood health index | -0.02 (-0.04, -0.003) | -1.3% * | 0.24 pp (0.01 pp, 0.47 pp) | -2.3% * |

| Trauma Sample |  |  |  |  |
| --- | --- | --- | --- | --- |
| Cognitive Score (0 - 27) |  |  | Cognitive Impairment |  |
|  | Coefficient (95% CI) | Percent Explained | Coefficient (95% CI) | Percent Explained |
| White | 15.5 (15.4, 15.6) |  | 18.4 pp (17.4 pp, 19.4 pp) |  |
| Black (reference) | 13.7 (13.3, 14.0) |  | 30.6 pp (27.5 pp, 33.7 pp) |  |
| Total gap between White and Black participants | 1.8 (1.5, 2.2) |  | -12.2 pp (-15.4 pp, -8.9 pp) |  |
| Gap explained by early-life circumstances | 0.74 (0.30, 1.2) | 40.4% ** | -7.9 pp (-12.0 pp, -3.9 pp) | 65.3% *** |
| Early-Life Circumstances (Variable-Level Decomposition) |  |  |  |  |
| Age | -0.41 (-0.58, -0.23) |  | 3.0 pp (1.4 pp, 4.6 pp) |  |
| Male | -0.05 (-0.11, -0.005) |  | 0.37 pp (-0.05 pp, 0.80 pp) |  |
| Marital status | 0.17 (0.04, 0.29) |  | -1.1 pp (-2.3 pp, 0.05 pp) |  |
| Born during WW2 | 0.03 (-0.01, 0.07) | 1.6% | -0.22 pp (-0.54 pp, 0.09 pp) | 1.8% |
| Born during Great Depression | -0.02 (-0.07, 0.03) | -1.0% | 0.16 pp (-0.26 pp, 0.58 pp) | -1.3% |
| Born in the South | 0.09 (-0.21, 0.39) | 4.9% | -0.75 pp (-3.7 pp, 2.2 pp) | 6.2% |
| Born outside of the US | -0.02 (-0.06, 0.02) | -1.1% | 0.23 pp (-0.19 pp, 0.66 pp) | -1.9% |
| Household size at age 10 | 0.02 (-0.10, 0.14) | 1.2% | -0.88 pp (-2.0 pp, 0.22 pp) | 7.2% |
| Relocated due to financial difficulties before age 16 | -0.01 (-0.04, 0.01) | -0.69% | 0.08 pp (-0.14 pp, 0.29 pp) | -0.63% |
| Family received financial help before age 16 | 0.03 (-0.01, 0.07) | 1.7% | -0.33 pp (-0.73 pp, 0.07 pp) | 2.7% |
| Father unemployed before age 16 | -0.0002 (-0.003, 0.003) | -0.01% | 0.002 pp (-0.04 pp, 0.04 pp) | -0.02% |
| Father worked a white collar job | -0.0006 (-0.11, 0.11) | -0.03% | -0.55 pp (-1.6 pp, 0.52 pp) | 4.5% |
| Mother did not work at age 18 | 0.04 (-0.12, 0.20) | 2.2% | -1.6 pp (-3.1 pp, -0.11 pp) | 13.1% * |
| Parental education (grouped) | 0.09 (-0.10, 0.28) | 4.9% | -0.69 pp (-2.5 pp, 1.1 pp) | 5.7% |
| Mother's education (years) | 0.11 (-0.05, 0.28) | 6.2% | -0.74 pp (-2.3 pp, 0.85 pp ) | 6.1% |
| Father's education (years) | -0.02 (-0.23, 0.19) | -1.2% | 0.06 pp (-2.0 pp, 2.1 pp ) | -0.47% |
| Education (years) | 0.42 (0.28, 0.56) | 22.8% *** | -3.3 pp (-4.5 pp, -2.0 pp) | 26.9% *** |
| Owned books at age 10 | 0.10 (0.003, 0.20) | 5.6% * | -0.72 pp (-1.6 pp, 0.18 pp) | 5.9% |
| Childhood health index | 0.0001 (-0.01, 0.01) | 0.01% | -0.0005 pp (-0.06 pp, 0.06 pp) | 0.004% |
| Childhood trauma index (EFTF) | 0.01 (-0.02, 0.04) | 0.51% | 0.05 pp (-0.22 pp, 0.33 pp) | -0.44% |
| Childhood trauma index (LHMS) | -0.03 (-0.15, 0.08) | -1.9% | 0.52 pp (-0.58 pp, 1.6 pp) | -4.3% |
| Relationship with mother before age 18 | -0.001 (-0.01, 0.01) | -0.07% | -0.02 pp (-0.12 pp, 0.08 pp) | 0.17% |
| Relationship with father before age 18 | -0.003 (-0.02, 0.02) | -0.18% | 0.04 pp (-0.18 pp, 0.26 pp) | -0.33% |

PGS Sample
| *Cognitive Score (0 - 27)* |  |  | *Cognitive Impairment* |  |  |
| --- | --- | --- | --- | --- | --- |
|  | Coefficient (95% CI) | Percent Explained |  | Coefficient (95% CI) | Percent Explained |
| White | 15.5 (15.3, 15.6) |  |  | 18.8 pp (17.7 pp, 19.8 pp) |  |
| Black (reference) | 13.6 (13.3, 14.0) |  |  | 31.1 pp (27.6 pp, 34.7 pp) |  |
| Total gap between White and Black participants | 1.8 (1.4, 2.2) |  |  | -12.4 pp (-16.1 pp, -8.7 pp) |  |
| Gap explained by early-life circumstances | 0.67 (0.11, 1.2) | 36.9% * |  | -6.1 pp (-11.5 pp, -0.80 pp) | 49.5% * |
| **Early-Life Circumstances (Variable-Level Decomposition)** |  |  |  |  |  |
| Age | -0.46 (-0.67, -0.25) |  |  | 2.9 pp (1.5 pp, 4.4 pp) |  |
| Male | -0.05 (-0.16, 0.07) |  |  | 0.09 pp (-0.80 pp, 0.98 pp) |  |
| Marital status | 0.20 (0.05, 0.35) |  |  | -1.4 pp (-2.8 pp, -0.08 pp) |  |
| Born during WW2 | 0.02 (-0.01, 0.06) | 1.2% |  | -0.15 pp (-0.41 pp, 0.12 pp) | 1.2% |
| Born during Great Depression | -0.02 (-0.09, 0.04) | -1.4% |  | 0.22 pp (-0.29 pp, 0.72 pp) | -1.8% |
| Born in the South | 0.06 (-0.30, 0.43) | 3.3% |  | 0.08 pp (-2.9 pp, 3.0 pp) | -0.67% |
| Born outside of the US | -0.04 (-0.09, 0.01) | -2.1% |  | 0.33 pp (-0.14 pp, 0.79 pp) | -2.6% |
| Household size at age 10 | -0.01 (-0.14, 0.13) | -0.44% |  | -0.49 pp (-1.6 pp, 0.58 pp) | 3.9% |
| Relocated due to financial difficulties before age 16 | -0.02 (-0.06, 0.02) | -1.1% |  | 0.03 pp (-0.26 pp, 0.31 pp) | -0.21% |
| Family received financial help before age 16 | 0.04 (-0.02, 0.09) | 1.9% |  | -0.32 pp (-0.75 pp, 0.12 pp) | 2.6% |
| Father unemployed before age 16 | -0.001 (-0.01, 0.01) | -0.07% |  | 0.002 pp (-0.05 pp, 0.06 pp) | -0.02% |
| Father worked a white collar job | -0.04 (-0.17, 0.10) | -2.0% |  | -0.59 pp (-1.7 pp, 0.55 pp) | 4.8% |
| Mother did not work at age 18 | 0.01 (-0.18, 0.20) | 0.65% |  | -1.3 pp (-2.9 pp, 0.35 pp) | 10.2% |
| Parental education (grouped) | 0.13 (-0.13, 0.39) | 7.1% | -1.3 pp (-3.4 pp, 0.82 pp) | 10.3% |
| Mother's education (years) | 0.17 (-0.04, 0.38) | 9.4% | -0.91 pp (-2.7 pp, 0.83 pp ) | 7.4% |
| Father's education (years) | -0.04 (-0.32, 0.24) | -2.2% | -0.36 pp (-2.6 pp, 1.9 pp ) | 2.9% |
| Education (years) | 0.44 (0.28, 0.60) | 24.0% *** | -2.9 pp (-4.3 pp, -1.5 pp) | 23.4% *** |
| Owned books at age 10 | 0.09 (-0.03, 0.22) | 5.2% | -0.59 pp (-1.6 pp, 0.42 pp) | 4.8% |
| Childhood health index | -0.0001 (-0.003, 0.003) | -0.01% | -0.002 pp (-0.05 pp, 0.05 pp) | 0.02% |
| Childhood trauma index (EFTF) | 0.003 (-0.04, 0.05) | 0.15% | 0.20 pp (-0.17 pp, 0.56 pp) | -1.6% |
| Childhood trauma index (LHMS) | -0.02 (-0.16, 0.12) | -1.1% | 0.30 pp (-0.83 pp, 1.4 pp) | -2.4% |
| Relationship with mother before age 18 | -0.002 (-0.02, 0.01) | -0.13% | -0.03 pp (-0.15 pp, 0.10 pp) | 0.22% |
| Relationship with father before age 18 | -0.01 (-0.04, 0.02) | -0.49% | 0.08 pp (-0.15 pp, 0.31 pp) | -0.62% |
| Polygenic scores | 0.04 (-0.07, 0.15) | 2.1% | 0.24 pp (-0.79 pp, 1.3 pp) | -1.9% |

<sup>a</sup> For clear presentation of the results, we keep one decimal point for estimates equal to or greater than 1 (e.g. 2.8), and keep two decimal points or more (if necessary) for estimates less than 1 (e.g., 0.07, 0.002).
<sup>b</sup> \*, \*\*, and \*\*\* besides cells corresponds to $p < 0.05$ , $p < 0.01$ , and $p < 0.001$ , respectively.

